# Assessment of Radiation Protection Compliance in Conventional Radiology Unit: A Cross-Sectional Study from Ibn Tofail Hospital, Marrakech

**DOI:** 10.64898/2026.04.27.26351706

**Authors:** Ahmed Warghad, adil Arza, Mohamed Bouibrine, Mohamed Saoud

## Abstract

**Background:** Radiation protection is a fundamental component of diagnostic radiology to minimize occupational and patient exposure to ionizing radiation.

**Objective:** To assess compliance with radiation protection standards and identify key deficiencies in conventional radiology units.

**Methods:** A descriptive cross-sectional study was conducted in the radiology department of Ibn Tofail University Hospital, Marrakech. Twenty-five occupationally exposed radiology personnel were included using exhaustive sampling. Data were collected using structured observation grids and self-administered questionnaires to assess compliance with radiation protection standards.

**Results:** The study revealed major deficiencies, including absence of individual dosimetry, inadequate structural shielding, poor quality control procedures, and non-compliance with occupational safety standards.

**Conclusion:** Radiation protection practices remain suboptimal, requiring urgent institutional reinforcement of regulatory compliance and enforcement, monitoring systems, and training programs.

## 1. Introduction

Ionizing radiation has become an indispensable tool in medical imaging, enabling accurate diagnosis and guiding therapeutic interventions. However, its use carries inherent risks, including DNA damage, carcinogenesis, and other stochastic and deterministic effects on both patients and healthcare workers (Hall & Brenner, 2008a). The fundamental principles of radiation protection—justification, optimization, and dose limitation—form the cornerstone of safe radiological practice and are essential to minimize these risks while maintaining diagnostic quality (ICRP, 2007). Despite comprehensive international guidelines established by the International Commission on Radiological Protection (ICRP) and the International Atomic Energy Agency (IAEA), compliance with radiation protection standards remains inconsistent across diagnostic radiology facilities worldwide. Evidence from multicenter audits and international assessments has documented significant deficiencies, including inadequate use of personal protective equipment, insufficient dosimetry monitoring, poor adherence to patient protection protocols, and gaps in quality assurance procedures. Furthermore, the UNSCEAR 2008 report highlights substantial variability in radiation protection practices and emphasizes that inappropriate or non-optimized use of ionizing radiation continues to contribute unnecessarily to professional exposure. Several occupational studies in radiology have reported suboptimal adherence to radiation protection practices, with inconsistent use of personal dosimeters, variable shielding compliance, and incomplete pregnancy screening protocols across institutions. In resource-limited settings, these challenges are often compounded by a lack of trained personnel, aged facilities, insufficient regulatory oversight, and limited institutional support for radiation safety programs. Despite limited support from the national radiation protection regulatory authority (Decret-n-2-97-30, 1997). Preliminary observations at Ibn Tofail Hospital, Marrakech, revealed concerning patterns of non-compliance with established radiation protection standards, including absence of routine dosimetry, inadequate structural shielding, and gaps in safety training among radiology technicians. This study aims to assess radiation protection compliance in conventional radiology units at Ibn Tofail Hospital, identify specific deficiencies in current practices, and propose evidence-based corrective measures to enhance safety for both patients and healthcare workers. Understanding the current state of radiation protection practices is essential for developing targeted interventions that align with international best practices and improve the overall safety culture within the institution.

## 2. Materials and methods

### 2.1. Study design and setting

A descriptive cross-sectional study was conducted in Ibn Tofail hospital (University Hospital Mohamed VI) of Marrakech. Data were collected from 01 May 2009 to 01 July 2009. This study is reported in accordance with the Strengthening the Reporting of Observational Studies in Epidemiology (STROBE guidelines) (Von Elm et al., 2007).

### 2.2. Participants and sampling

An exhaustive sampling design was adopted to recruit only technicians as participants working with ionizing radiation in radiology unit. All radiology personnel occupationally exposed to ionizing radiation and actively working during the study period were included. Trainees, temporary staff were excluded. Twenty-five participants were included. Participation was voluntary, and only participants who provided informed consent. Compliance was assessed according to conformity with national radiation protection guidelines of radiation protection.

### 2.3. Data collection instrument

Data were collected using both an observation grid and self-administered paper questionnaire distributed face-to-face. The primary tool was used to assess structural indicators applied in radiology unit in accordance with local regulation (X-ray warning signage, architecture of room control console shielding, dosimetry etc…). The second instrument included contain socio-demographic characteristics (age, gender, specialty, date of affectation) also self-reported perceptions of radiation protection practices in the unit by technician for all items, which were cross-validated using the observation grid ensuring complementarity of data collection. The data used in this study were originally collected in 2009 as part of an academic institutional research project conducted during professional training at IFCS Marrakech. The present manuscript represents a retrospective scientific analysis of these archived data with the objective of documenting baseline radiation protection practices and contributing to current academic and professional development in radiological safety.Although data were collected in 2009, they provide a baseline assessment of radiation protection practices prior to recent regulatory reinforcement in Morocco.

### 2.5. Bias Control

Data were collected using a self-administered, paper-based questionnaire. Participants were guided during questionnaire completion to ensure comprehension of the items, reducing information bias. The use of both direct observation and self-reported measures enabled methodological triangulation, allowing comparison between observed compliance and reported practices, thereby improving internal validity and reducing single-method bias.

### 2.6. Data analysis

Data were entered, cleaned, and analyzed using statistical software PASW Statistics software. Data quality checks were performed. Descriptive statistics were used to summarize the data: Continuous variables were reported as mean ± SD or median (IQR), depending on distribution. Categorical variables were expressed as frequencies and percentages. Radiation protection compliance was analyzed using the observation grid and reported as proportions of compliance versus non-compliance for each safety indicator (e.g., shielding, signalization, room design).

### 2.7 Ethical approvals

This study involved human participants and questionnaire-based occupational data collection among radiology personnel. Ethical approval was granted by the Research Ethics Committee of the Institut Supérieur des Professions Infirmières et Techniques de Santé de Marrakech (The Higher Institute of Nursing Professions and Health Techniques of Marrakech), affiliated with the Ministry of Health and Social Protection, Morocco, under approval number No. 24/2009/IFCSM.

The committee reviewed and approved the study protocol, before initiation of data collection. Written informed consent was obtained from all participants prior to inclusion in the study. Participation was voluntary, and all collected data were anonymized and handled confidentially in accordance with institutional ethical standards and applicable national regulations.

## 3. Results

### 3.1. Sociodemographic Characteristic of study population

A total of 25 radiology personnel were included in the study, consisting of 17 radiology technicians (TRX), 5 general nurses (IP), and 3 application engineers. The study population comprised 15 females (60%) and 10 males (40%) predominantly young to middle-aged adults, with an age range of 22–40 years. A marked heterogeneity in occupational seniority was observed across professional categories. Radiology technicians reported 3–7 years of professional experience, while supervising professionals (IP) had more than 10 years of professional experience. Application engineers demonstrated the highest experience level, with approximately 12 years of seniority, reflecting long-term involvement in radiological work.

#### 3.2.1 Grid Results

#### 3.2.1 Structural and architectural compliance of radiology rooms. (Table 1)

Radiology room surfaces exceeded the recommended standard (28.5 m^2^ - 27.25–27.27 m^2^ vs 18 m^2^). However, overall architectural design was non-compliant with radiation protection requirements. Key deficiencies included inadequate structural separation of control consoles, lack of integrity verification of shielding elements (including broken lead glass and doors), failure to respect the required non-respect of required tube–console distance, suboptimal tube positioning, thereby increasing the risk of scattered radiation exposure, and absence of controlled area signage and warning systems.

**Table 1.** Architectural compliance of radiology rooms (observation grid results)

| Parameter | Observation | Standard / Reference | Compliance |
| --- | --- | --- | --- |
| Room surface area | 27.27 m<sup>2</sup> / 27.25 m<sup>2</sup> | 18 m<sup>2</sup> | compliant |
| Control console location | Inside room | Outside | Non-compliant |
| Lead shielding integrity | Not verified | Periodic testing required | Non-compliant |
| Tube–console distance | Not respected | maximum distance required | Non-compliant |
| Tube positioning | Suboptimal | Optimized required | Non-compliant |
| Warning signage | Absent Mandatory | controlled area signage | Non-compliant |

#### 3.2.2 Equipment-related radiation protection deficiencies. (Table 2)

All radiology installations (Italray system 1998), which had been installed for more than 10 years, were associated with recurrent monthly technical issues. Maintenance and quality control assessments revealed multiple deficiencies, including absence of routine calibration of photoelectric parameters, lack of tube output verification, absence of photogeometric quality, incomplete beam alignment checks, improper or absent anti-scatter grid alignment, and occasional use of malfunctioning diaphragms.

**Table 2.** Equipment quality control and radiological safety checks.

| Parameter | Status | Frequency expected | Compliance |
| --- | --- | --- | --- |
| Photoelectric calibration QC | Not performed | Routine QC | Non-compliant |
| Tube output verification | Not performed | After maintenance | Non-compliant |
| Photogeometric control | Not performed | Routine QC | Non-compliant |
| Beam alignment | Not verified | Routine QC | Non-compliant |
| Anti-scatter grid alignment | nconsistent | Routine QC | Non-compliant |
| Diaphragm function | Sometimes defective | Continuous check | Non-compliant |

#### 3.2.3. Occupational practices and staff-related compliance. (Table 3)

Multiple unsafe radiation protection practices were observed, including non-optimized fluoroscopy time use, frequent exposure due to open doors during imaging, absence of beam collimation devices, lack of protective equipment and aids, failure to maintain the required Source-to-patient distance (SPD), absence of personal dosimetry, and lack of patient immobilization devices.

**Table 3.**
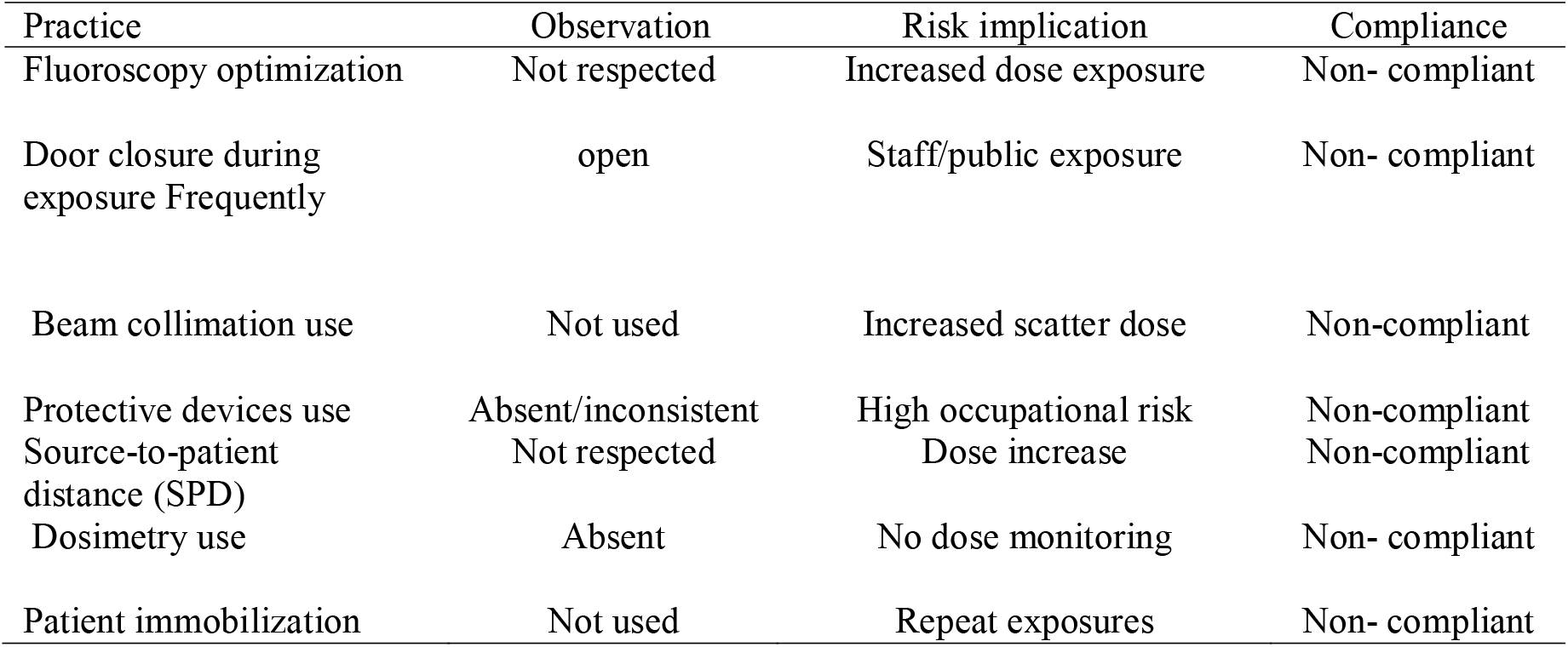
Occupational radiation protection practices.

### 3.5. Questionnaire results (table 4)

All participants (100%) reported knowledge of radiation protection standards 64 % through basic training and 32% as continuous training related to their professional role in the unit. 83.3% of technicians reported the presence of deficiencies in radiation protection measures : 91.6% reported absence of controlled/supervised area signage, 88 % judged the architectural design inadequate for radiation protection, 100% reported absence of individual dosimetric monitoring, 88% reported absence of regulatory quality control, 100% reported absence of systematic monitoring of radiation output after maintenance, 92% considered shielding ineffective or unreliable. 84% reported absence of protective lead aprons,100% reported absence of periodic occupational medical examinations. In summary 88 % of technicians reported dissatisfaction with radiation protection conditions within the department.

**Table 4.** Results of questionnaire:

| Variable | Category | Count/number | percentage % |
| --- | --- | --- | --- |
| Knowledge of RDP | Yes | 25 | 100 |
| Training type | Basic training | 16 | 64 |
| Training type | Continuing training | 08 | 32 |
| Training type | Self-learning | 01 | 04 |
| Perceived deficiencies | Yes | 20 | 83.3 |
| Warning signage present(zoning) | No | 4 | 91.6 |
| Architectural adequacy | Not adequate | 22 | 88 |
| Dosimetry monitoring | No | 25 | 100 |
| Quality control (NCRP/PCR) | No | 22 | 88 |
| Tube radiation control | No | 25 | 100 |
| Lead glass reliability (shielding) | No | 23 | 92 |
| Lead aprons availability | Absent | 21 | 84 |
| Medical surveillance | No | 25 | 100 |
| Overall protection radiation satisfaction | Not satisfied | 22 | 88 |

## 4. Discussion

The present study demonstrates systemic deficiencies in radiation protection compliance in equipment quality control and occupational practices within conventional radiology units with respect to national regulatory requirements (Decret-n-2-97-30, 1997). These findings are consistent with international evidence indicating that non-compliance with radiation safety standards was already a global concern. The absence of individual dosimetric monitoring observed in this study represents a violation of radiation protection principles. Previous literature emphasizes that individual dose monitoring is an essential component of radiation protection programs, yet remains insufficiently implemented in some medical settings, leading to uncertainties in the assessment of occupational exposure (Vanhavere et al., 2008). From a radiobiological standpoint, ionizing radiation is well established to induce stochastic effects, including carcinogenesis, through DNA damage and mutagenesis mechanisms (David Brenner & Eric Hall, 2007). The absence of systematic individual occupational dose monitoring constitutes a critical issue of radiation protection practice, which is an essential component of dose limitation and optimization principles (ICRP, 2007; Filip Vanhavere et al., 2008). The absence of dosimetric follow-up prevents accurate assessment of cumulative exposure and undermines effective risk management for healthcare workers (ICRP, 2007). In parallel, quality control procedures including tube output verification, beam alignment, and post-maintenance calibration are essential to ensure accurate and reproducible radiation delivery. Any failure of these processes leads to unpredictable and unintended patient exposure due to technical inaccuracies. Such deficiencies constitute a violation of the optimization principle as defined by ICRP Publication 103 (ICRP, 2007). Structural deficiencies identified in this study, including poorly maintained shielding elements (including damaged lead glass and doors) and absence of controlled area zoning and the absence of exposure warning systems reflect non-compliance with established radiation protection infrastructure standards. These elements are fundamental requirements in international radiological protection frameworks (IAEA Safety Standards (GSR Part 3)). Moreover, unsafe occupational practices such as failure to use collimation, improper positioning, and lack of protective equipment are consistent with earlier evidence demonstrating increased operator and patient exposure and reduced image quality due to suboptimal techniques or alternative inadequate techniques contributing to increased radiation exposure for both patients and operators due to suboptimal technique optimization.(ICRP 2007). Notably, despite universal self-reported knowledge of radiation protection among participants, although universal self-reported knowledge was observed, compliance remained poor, this persistent gap between theoretical knowledge of radiation protection and clinical compliance has been reported in radiology practice, suggesting that system-level and organizational factors may influence adherence to radiation safety principles more than individual awareness. As limitations of our study, the sample size was limited and the study is single-center, the findings remain internally consistent. Although the cross-sectional design precludes causal inference, it provides a reliable snapshot of real-world radiation protection practices. Potential self-reporting of many items regarding the experience of working conditions of working with ionization radiation may have been affected by the subjectivity of participants was addressed by methodological triangulation. The lack of quantitative dosimetric measurements for exposure estimation and the absence of longitudinal worker follow-up should be acknowledged. Overall, these findings reflect deficiencies in the implementation of the core principles of justification, optimization, and dose limitation, particularly in settings with limited regulatory oversight.

The absence of dosimetry, inadequate quality control and unsafe occupational practices represent major violations of ICRP recommendations. This is consistent with international reports highlighting that non-optimized use of ionizing radiation contributes significantly to avoidable exposure (UNSCEAR, 2008) Radiation protection practices are suboptimal. Immediate corrective actions are therefore required to ensure safety and compliance. An effective radiological protection system requires mandatory individual dosimetric monitoring to ensure accurate occupational dose tracking and compliance. Governance should be reinforced through a designated Radiation Protection Officer (RPO) overseeing safety implementation and audits. A structured quality assurance program must include routine verification of beam output, alignment, and calibration. Facilities should ensure compliant shielding and controlled area classification. Continuous competency-based radiation safety training is essential, alongside strict enforcement of personal protective equipment (PPE) use. Collectively, these measures optimize exposure control under the ALARA principle and strengthen the overall culture of radiological safety.

## Data Availability

Data were collected in the cadre of final report of Obtaining licence in ISPITS/IFCS OF MARRAKECH;

## BIBLIOGRAPHIE

Décret n° 2-97-30 du 25 joumada II 1418 (28 octobre 1997) pris pour l’application de la loi n° 005–71 du 21 chaabane 1391 (12 octobre 1971) relative à la protection contre les rayonnements).

Dendy, P. P. (2008). Radiation risks in interventional radiology. The British Journal of Radiology, 81(961), 1–7. 10.1259/bjr/15413265 Hall, E. J., & Brenner, D. J. (2008). Cancer risks from diagnostic radiology.

The British Journal of Radiology, 81(965), 362–378. 10.1259/bjr/01948454 ICRP, 2007. (2007). Preface, Executive Summary and Glossary. Annals of the ICRP, 37(2-4), 9–34. 10.1016/j.icrp.2007.10.003 Kuipers, G., Velders, X. L., De Winter, R. J., Reekers, J. A., & Piek, J. J. (2008).

Evaluation of the Occupational Doses of Interventional Radiologists. CardioVascular and Interventional Radiology, 31(3), 483–489. 10.1007/s00270-008-9307-7

Malone, J. F. (2009). Radiation protection in medicine□: Ethical framework revisited. Radiation Protection Dosimetry, 135(2), 71–78. 10.1093/rpd/ncp010

Rehani, M. M. (2008). The IAEA’s activities in radiological protection in digital imaging. Radiation Protection Dosimetry, 129(1-3), 22–28. 10.1093/rpd/ncn155

United Nations Scientific Committee on the Effects of Atomic Radiation. (2010). Sources and Effects of Ionizing Radiation, United Nations Scientific Committee on the Effects of Atomic Radiation (UNSCEAR) 2008 Report, Volume I□: Report to the General Assembly, with Scientific Annexes A and B - Sources. UN. 10.18356/cb7b6e26-en

Vanhavere, F., Carinou, E., Donadille, L., Ginjaume, M., Jankowski, J., Rimpler, A., & Sans Merce, M. (2008). An overview on extremity dosimetry in medical applications. Radiation Protection Dosimetry, 129(1-3), 350–355. 10.1093/rpd/ncn149

Von Elm, E., Altman, D. G., Egger, M., Pocock, S. J., Gøtzsche, P. C., Vandenbroucke, J. P., & for the STROBE Initiative. (2007). The Strengthening the Reporting of Observational Studies in Epidemiology (STROBE) Statement□: Guidelines for Reporting Observational Studies. Annals of Internal Medicine, 147(8), 573–577. 10.7326/0003-4819-147-8-200710160-00010

